# Development and Iterative Design of Peer-based Discussions about Heart Health for Women

**DOI:** 10.1101/2024.05.09.24307138

**Authors:** Erica S Spatz, Tina Park, Maggie Breslin

## Abstract

**Objective:** To support women in their health journeys around cardiovascular risk reduction, providing: access to information; space to discuss complex and personal issues that may factor into decision making; and skills to support conversations with healthcare providers.

**Methods:** We used a participatory design approach to develop peer sessions led by a non-clinical facilitator and a cardiologist, for women to learn, think, and discuss information and decision-making around cardiovascular risk. These sessions took place in-person and over video. A host invited her friends, family or network to the events. Sessions were designed to bring information to women at all stages of cardiovascular risk, including women with no prior experiences with heart health and women with established cardiovascular disease. The sessions provided information about cardiovascular risk and facilitated discussion around risk factors and treatment. Sessions were recorded, transcribed and analyzed using a grounded theory approach to identify emerging themes.

**Results:** We conducted 12 groups, with an average of 6 women per group. Women were of all ages and spectrums of cardiovascular risk. The three major themes that emerged were: Starting Place (attitudes, beliefs, and behaviors toward cardiovascular care are informed by a blend of one’s family history, personal interactions with their health and the health system, and approach to managing uncertainty); Lack of Sense-Making (seemingly simple recommendations to support heart health were much more complicated when related to their own risk and experience), and Self-Judgement and Judgement by Others (without support or validation, women may feel judged by others for not doing enough).

**Conclusion:** Peer sessions can illuminate the complicated issues women face when approaching decisions regarding cardiovascular health. These sessions may offer an alternative to the healthcare setting, for women to wrestle with complex issues that relate to their risk, and better prepare for visits with their healthcare providers.

**Highlights:**

- Peer-based sessions designed to support cardiovascular risk reduction conversations can support women in their healthcare journeys.
- Women approach cardiovascular risk from different starting places and are challenged with sense-making of conflicting data.
- Women are often judged by themselves, and others, about doing ‘enough’ to prevent cardiovascular disease.
- Peer-based sessions offer a space for women to think, feel, and discuss issues most relevant to them, which may help support conversations with their healthcare providers.

---

Despite decades of evidence that cardiovascular risk in women is frequently underestimated and that symptoms of heart disease are often dismissed, and despite public health efforts to bring more awareness to women’s heart disease, care models have not evolved to better engage women in their heart health. In the standard medical encounter for both men and women, conversations are centered around the traditional cardiovascular risk factors-including blood pressure, glucose, smoking and cholesterol – data which are readily available and easily quantified in the electronic health record, and can be input into calculators to estimate individuals’ future risk of heart disease. Other biological factors that are relevant in women such as pregnancy outcomes, menopause, inflammatory disease and family history, along with lifestyle factors such as stress, diet and exercise – all of which can modify risk, are infrequently evaluated or discussed.^1^ Moreover, women may be unaware of the importance of these factors in assessing and managing cardiovascular risk and may find it challenging to raise these issues during the clinical encounter.^2^ Yet these factors more fully capture a woman’s biology and biography, her so-called “lived experience,” and are essential components of cardiovascular risk assessment and management.

Drawing from our considerable experience developing shared decision making (SDM) tools for patients and clinicians to use in the encounter,^3-7^ we hypothesized that an intervention for patients that helped build their skills and confidence in articulating and then sharing personal information relative to their cardiovascular risk with their clinician might be a meaningful companion program. We imagined combining a trusted source for information about cardiovascular disease and prevention with a space for patients to identify the stories from their lived experience that are most meaningful to managing their heart health. We envisioned peer groups and a setting outside of the clinical visit. We set out to design and iteratively develop these sessions with prospective participants to meet the goals and expectations of diverse women with and without established cardiovascular disease.

Accordingly, we developed a group session facilitated by healthcare designers and including a preventative cardiologist to engage women in a peer-based cardiovascular risk communication session with an emphasis on the elements of “biography” that are so often missing from the clinical encounter. The purpose of these sessions was to create a space for women to think, feel, and talk about the challenges of cardiovascular disease prevention, identify areas that were important to them, and gain confidence in sharing their stories and questions with their doctors. In this paper we describe the development of the peer sessions and the key themes that emerged from qualitative analysis of the sessions. It is our hope that this information can be used by other clinicians and designers working together to develop programs to support women in their cardiovascular health journey.

## METHODS

We used a participatory design approach based on principles from human-centered design, and conducted a qualitative analysis using grounded theory, to incorporate end-user feedback and identify key themes that would ensure the event, facilitator guidebook and patient-facing materials were usable, accessible, and effective. We first synthesized data on cardiovascular disease risk in women by examining the literature for what is already known about women’s knowledge, preferences and concerns about cardiovascular disease prevention. With this foundation, and using our own clinical, professional, and research experience, we developed the initial peer session and materials. Based on participant feedback and analysis of audio-recorded transcripts from the peer sessions, we iteratively developed the peer sessions and materials, until we felt we were achieving consistent conversational results regardless of setting (in person or on Zoom), that we’d identified the key activities and necessary participants for a successful session, and that the materials developed were suitable for individuals in different stages of life and with different cardiovascular health backgrounds, levels of knowledge, and comfort with engagement.

### Development of Peer Group Sessions

The peer groups served as both an intervention and a lab for further developing the sessions and tools. The peer group events were co-developed by cardiologists and designers to:

- Enhance the interest and comfort levels of women from different age groups, levels of risk and experience with the healthcare system in engaging in a group session about cardiovascular health
- Provide information about cardiovascular disease and standard tools used in the clinical setting to assess individuals’ risk of having a future heart attack.
- Introduce the concept of one’s biology and biography influencing their cardiovascular health, and to discuss sex-specific issues that may often be overlooked in these discussions
- Discuss the role of statins in reducing cardiovascular risk and create better understanding about how the benefits of a statin change depending on a woman’s baseline risk; information on calcium scoring was also introduced as a screening tool for cardiovascular disease and as a tie breaker if conflicted about whether to take a statin. In these discussions we explored how a woman’s preferences, values and goals may influence her decision to take a statin or go forward with a coronary artery calcium score.
- Role play conversations between physicians and patients about the decision to take a statin, and see how these concepts can be introduced during the conversation
- Review what women took away from the sessions and how this information and experience may influence their next conversation.

In planning each session, one participant was asked to host – either at her home or another location. When Covid-19 disrupted in-person gathering, we moved to facilitating the events on zoom. The host invited women in her social network, through work and word of mouth to attend. Approximately 5-12 women participated in each session. Two additional sessions were structured to specifically capture women of color, who were underrepresented in the initial groups. Each session lasted about 90 minutes and was structured such that the facilitator (healthcare designer, MB) led the group in discussion about what came to mind when they thought about their heart health, asked them to reflect on previous conversations with their clinician about heart health, and introduced the biology and biography framing for risk factors of cardiovascular disease **(Figure 1)**. The cardiologist, trained in patient engagement and shared decision making, then discussed the sex-specific factors that play a role, inviting conversation and questions along the way. The facilitator then described the Mayo Statin Decision Aid and walked through 3 scenarios using the aid.^8-10^ Finally, a second designer (TP) and the physician role-played the 3 scenarios to highlight the ways that women with similar risk profiles but different contexts can have different conversations about their heart health and make different decisions about what to prioritize. A booklet was created to support the in-person sessions and elements of the booklet were adapted to support the event over zoom **(Supplement 1)**. Following each session, we invited informal (not anonymous) and formal (survey-based; anonymous) feedback; unfortunately, we had very few surveys returned and cannot include these data as part of the evaluation. In total, 12 sessions were conducted with 93 women.

**Figure 1.**
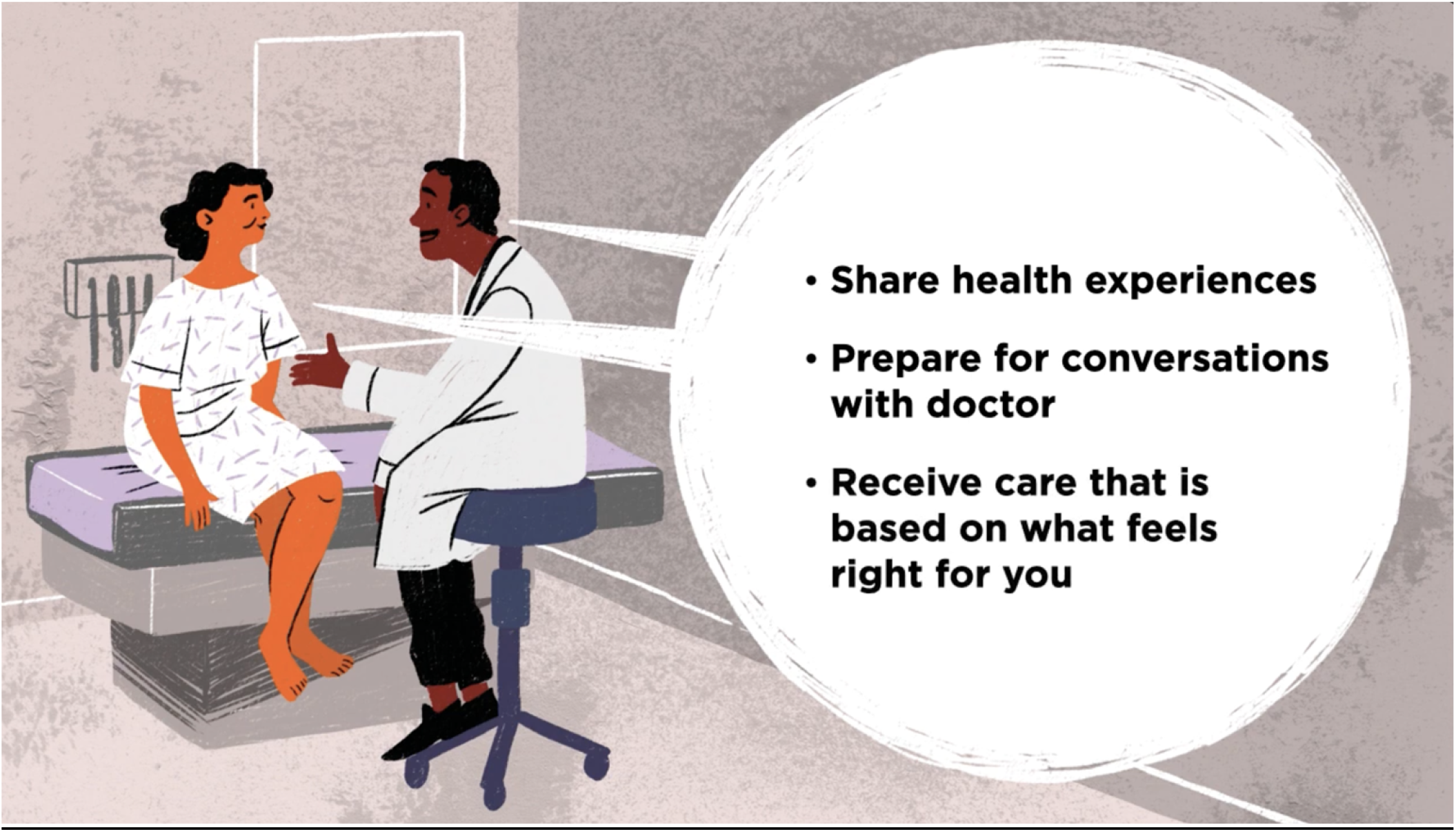
The sessions were introduced by a brief [video](https://www.youtube.com/watch?v=_PBLJfhFKSg), highlighting what to expect from the session as well as the goals of the sessions: to share health experiences, prepare for conversations with their doctor, and to promote care that feels right for them.

### Qualitative Analysis

All events were audio recorded and 8 of the 12 sessions were of high enough quality to allow for post-event analysis; 4 conducted in person and 4 conducted over zoom. For the thematic analysis, we used an inductive approach, allowing the themes to emerge from the interview transcripts. We used an iterative process to develop the code structure. Initial codes were identified by group reflection, then evolved based on transcript review with the same reviewers assigned to transcripts. Once codes were defined, two reviewers independently coded each transcript. Emergent themes are described in the Results section.

The study was approved by the Yale Human Investigations Committee with verbal consent by participants obtained by ESS and MB, and witnessed by TP. The study was funded by the Alpha Phi Foundation, Heart to Heart Grant.

## Results

Participants were recruited from March 1, 2019 through August 30, 2021. The mean number of attendees were 7 (range 5 to 14). Attendees were diverse in terms of age and prior experience with cardiovascular disease. Although we did not collect data about women’s sociodemographic or medical histories, we did observe that some groups had women spanning young adulthood to mid- and late-adulthood; while other groups were more similar in their ages. During the sessions we learned that the women within each group had very different medical backgrounds, ranging from no medical history to having already had a heart attack or other heart condition, from being very committed to cardiovascular health to not wanting to engage much, and from having no prior conversations with their physicians to having several cardiologists involved in their care. Many women were already taking statins. A priori, some women expressed skepticism or hesitation about statin medications and other women felt very strongly that statins were important and should be used more often.

With the thematic analysis, we sought to identify common issues that women brought up and seemed to be grappling with on their own, in their family, and with their care team. Several themes emerged from the sessions **(Figure 2)**. They included:

**Figure 2.**
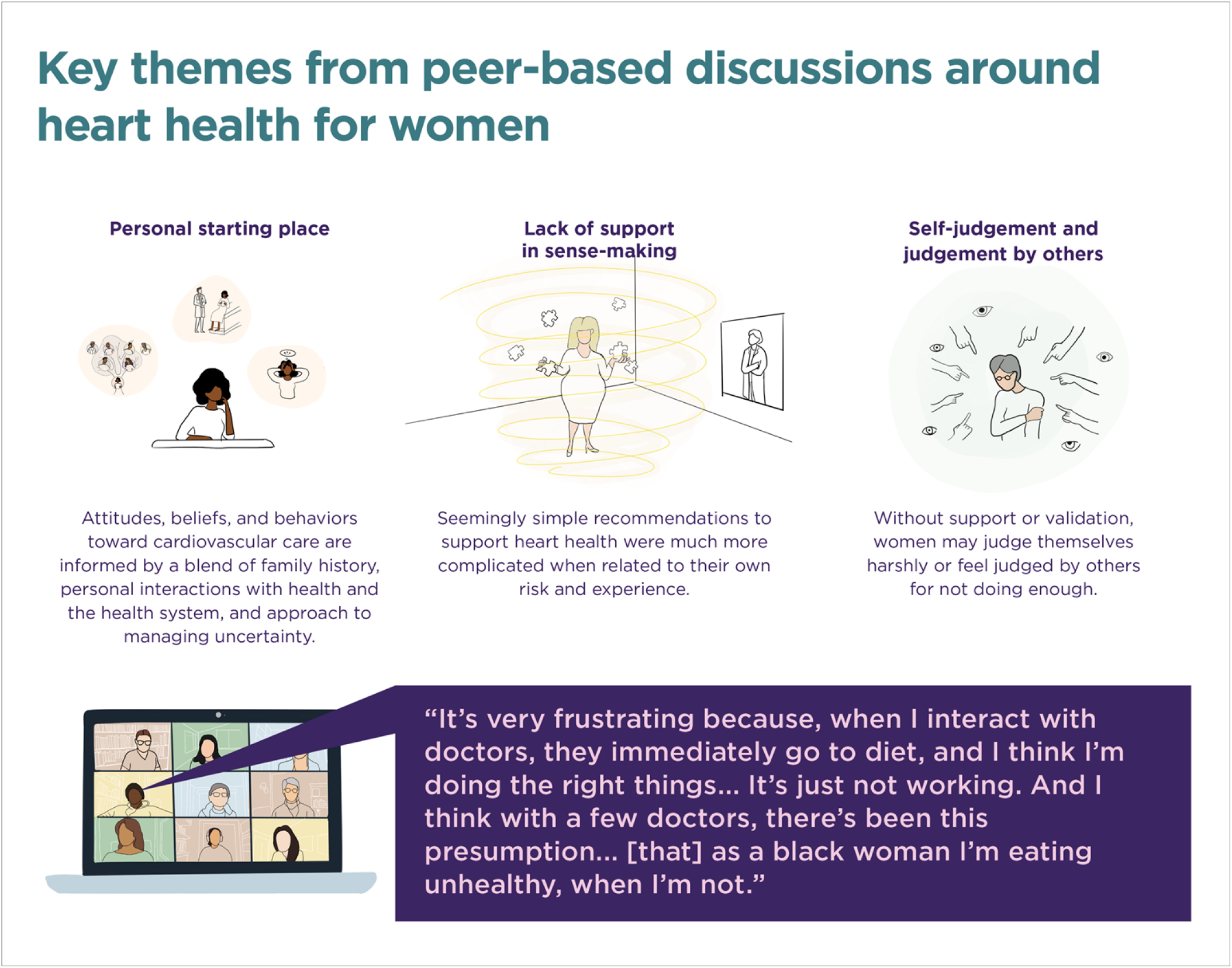
Emergent themes from 8 peer sessions with women to discuss heart health.

### Starting Place

Most/all women in our events were familiar with the traditional risk factors of diet and exercise, smoking, high blood pressure, high cholesterol, and diabetes. What was unique to each woman was how they blended that information with their own family history, their personal interactions with their health and the health system, and their approach to managing uncertainty. This active synthesis of the personal and the common affected a woman’s attitudes, beliefs, behaviors, and overall engagement with cardiovascular information and care.

The opening question of the heart health events – “when you think about your heart health, what comes to mind?” offered an opportunity for each woman to identify the issues and concerns that were top of mind for themselves, and also to hear how other women processed similar information with their own experiences and arrived at a different set of concerns and issues. A mother and daughter who participated in one of our sessions had very different responses to this question even though they share a genetic mutation that increases their cardiovascular risk.

> *“For me, when I think about heart health, I don’t. I don’t think about it very often. It’s just more like, I think it’s brought up when I’m around my family. It’s like, oh, you’re eating bacon. Oh, you’re eating fries. You’re not supposed to eat that. You should do this. Because it’s very common conversation in our family. But it’s one that I kind of like to avoid, because I don’t, I don’t do all the things that you’re supposed to do. Because I feel like I just don’t really want to. Maybe I’ll deal with that like in my 30s or my 40s*.*” (daughter)*

> *“When I think about heart health, it’s a very stressful thought for me. I mean, it’s, it’s every day it’s you know, did I remember to take my statin and is my statin working and am I eating okay, am I exercising enough. And, you know, we’ve had two of our brothers have heart attacks. And, you know, my mother, I think had what they called silent heart attacks, which is one of my questions for later. So, you know, I’m constantly worrying, you know, I’m 50, almost 50 and just, you know, am I going to have a heart attack? I just want to get more information and get a better handle on what I need to do*.*” (mother)*

### Lack of support in sense-making

Cardiovascular risk prevention advice often boils down to simple recommendations; *exercise for at least 30 minutes 5 days a week, reduce your stress, go to the emergency department immediately if you are having chest pains*. However, we observed in these sessions that it can be challenging for women to figure out how to integrate this advice into their complex lives and they are mostly left on their own to do it. Additionally, integrating recommendations about lifestyle and genetic/biological factors to reduce their own cardiovascular risk can be confusing, especially when they are managing multiple chronic conditions.

> *“So he was among the cardiologists that send me to the dietician. But the dietitian, although (the cardiologist) sent me, is not focusing on the heart, they is focusing on the sugar. So what she want is my A1c3 to go down. So she put me on very low 90 carbs a day. Uh huh. So, you know, I mean, I cry, but how do I balance? Right? What I should not have to protect my heart? And what should I not have so my sugar doesn’t go up?”*

They also expressed confusion about what to do when they had heart symptoms, not wanting to ignore symptoms that may be concerning, but also feeling dismissed by healthcare providers when raise concerns or go to the emergency department. For example, one woman described having chest pains that were concerning enough for the doctor to recommend she seek emergency medical care, but when the tests returned normal, her symptoms were never attended to and there was no continuing effort to figure out what was wrong. She reported feeling conflicted, knowing women can having different symptoms than men, but also not knowing when to seek care.

> *“When you call your doctor and say, you’re having chest pain, their immediate response is go to the emergency room, which I did. And if you go to the emergency room and say you’re having chest pain, they run a boatload of tests…I was pretty young still. But as someone who’s in public health after having that experience, I’m reluctant to do that, again. Because I’m also aware of health care spending, and not wanting to overuse healthcare resources. So that puts me in all kinds of binds, right? Because I never know what to do…, when I am having a heart issue… does that warrant emergency attention, or not. So as a woman, I think it’s particularly complicated, because I know that symptoms are very different in women, and I don’t always necessarily know how, what that entails*.*”*

### Self-judgement and judgement by others

Judgment, in a number of forms, was a common theme in the event discussions.

Without support or validation, women may judge themselves harshly or feel judged by others for not doing enough.

### Self-judgment

Some women in the groups who were either considering or were already taking preventive cardiovascular medication tended to blame themselves for needing the medication. They spoke about knowing they should eat more healthily, or exercise more, or lose weight, but couldn’t ultimately “do enough” to improve their blood pressure or cholesterol. Some reported stress to be a major factor in their lifestyle behaviors.

> *“… I’ve been on insulin forever and it didn’t really feel like that big of a deal to add another pill. But I will say that, like, every night when I’m taking my statin … there is some I don’t know, disappointment about like, Oh, I couldn’t, you know, do this myself, or what does it mean to be this young and be on a statin, and kind of all those things and it’s just that you know, that small kind of like, nag of like, Oh, yeah, maybe I could be doing this better myself. …what does that decision say about me and my ability to care for myself?”*

### Feeling judged by others, including clinicians

Some women expressed feeling judged by their doctors. Women of color expressed bias in their healthcare interactions. For example, women repeatedly heard messages that they were not exercising or dieting enough, and comments that had they been following recommendations, their numbers might have come down. Women also reported that they perceived their doctors as frustrated and dismissive when there was uncertainty about a diagnosis, even though women were not expecting them to have all of the answers.

> *“But it’s very frustrating because, you know, when I interact with doctors, it’s, you know, they immediately go to diet, and I think I’m doing the right things. It’s just not working. And I think with a few doctors, there’s been this presumption, I think as a Black woman [they think] I’m eating unhealthy, when I’m not. And I am exercising. But it’s very frustrating and I worry a lot about my heart health. I have a sister also with cardiovascular disease. So I worry about my siblings and my children. Very, very stressful*.*”*

### Nature of cardiovascular advice

Common cardiovascular advice is generic, plentiful, and intimately tied to everyday activities, often leaving women with both the knowledge and the feeling that there is more they could be doing to improve their heart health.

> *“… exercise more, And don’t drink, cut out that wine and of course, the high cholesterol rule. And that’s it? I did have a lot of stress. And he just said basically something like you know, we all have stress, we have to learn how to deal with it. You should try meditation or something like that. Okay, what else moving on*.*”*
>
> *“diet, eat less sugar, eat less. which of course I don’t do*.*”*
>
> *“Yeah, I’m trying with the diet, it’s hard to find time…, you know, My motivations have just gone way down. And I like, I think about it, and I know that I should, but I probably I should be doing better*.*”*
>
> *“I don’t know whether I should be taking statins. And I know there are side effects. I’d like not to take drugs. I yeah, I don’t take drugs. And I don’t want to start. I do exercise. (Participant 1) will tell you I eat a lot of broccoli*.*”*

### Integration of themes into facilitators guide and patient materials

These themes helped us evaluate and prioritize activities and information within the sessions so we could create a safe and welcoming space for the type of sense-making that was emerging. Additionally, the themes revealed shared issues and anxieties among women that impact their cardiovascular health that may also be present in the management of other conditions. And finally, the themes offer an initial set of heuristics to use in considering how these sessions are different from other educational/informational sessions and lectures which are more typical of health events.

## DISCUSSION

Throughout a woman’s lifecourse there is a need for new levels of awareness, knowledge, and support about cardiovascular health. Information about cardiovascular health is often restricted to the clinical encounter, or to educational materials or sessions. Rarely are women given the opportunity to react to information by experts and consider how the information applies to their own life. Additionally, there are no forums for women to think, feel, and talk about the information, and how they are making decisions. We learned from these sessions that there is a need for this type of interaction.

Sessions were conducted with peers which facilitated openness – as there was often a baseline of trust and/or familiarity with one another in the group. We found that women often shared divergent experiences and opinions about the data or about their own cardiovascular care, but that they learned from one another and gained a broader sense for how others approached these decisions. We also learned that women often do not have the opportunity to think through these issues ahead of their visit with their medical provider and are therefore unprepared to talk about the issues that are most relevant to them or to ask questions about what is right for them. For example, there was great interest in stress and its relationship to cardiovascular disease, a desire to understand more about how statins work – beyond their effects in lowering cholesterol, and how a coronary artery calcium score may help them to understand more about their cardiovascular risk and whether statins are right for them. Many women had a chance to see the patterns of healthcare interactions that made them feel judged by their clinicians, or to judge themselves, and were able to ask questions about why this was the case. A common theme was whether diet and exercise were truly effective in lowering numbers, and whether having a family history of early heart disease was sufficient to put a person at risk for having a heart attack.

These questions are critical to the health belief model, wherein for behavior change to occur a person must understand their risk, perceive that they are at elevated risk, and feel capable of changing that risk.^11^ Moreover, the interventions need to be acceptable. For many women previously reluctant to take a statin, they heard from “like” women about why they chose to take one, and reported being more open to it in the future. Many women realized that they didn’t know their risk and were going to make it a point to discuss at their next doctor’s visit. Some women, especially those who had cardiovascular disease, gained a more nuanced understanding of their condition and identified with new ways that they could improve their lifestyle and care interactions.

There are several limitations that are important to address. One is that in this type of work, it is challenging to capture effectiveness. One is that there is no agreed upon measure for when a one-time peer intervention is effective. Our goal was not to improve an outcome, but rather to test the hypothesis that women were interested in engaging and that the sessions were meaningful. Larger studies could assess whether the sessions facilitated higher quality patient-clinician interactions, or changed lifestyle behaviors or care decisions. This was beyond the scope of this project. Another limitation is that we did not collect specific sociodemographic factors, which ended up to be more challenging than we planned. Although we attempted to conduct groups with a diverse group of women, it was difficult to know apriori who would come to the events; many women ‘showed up’ for the host, as opposed to coming for the purpose of the information. However, we purposefully sought out groups of different ages, races, and ethnicities. One event was with mostly young women; another event was with retirees in a book club. Another event was intergenerational with a family. Some groups included women of different races; some groups included only women who were Black or white, which mostly reflected the hosts’ peers. More purposeful testing with groups of different ages, health status, race and ethnicity can help to ensure that the events are appropriate for all groups. Finally, obtaining responses to surveys was challenging, and most people did not fill out a brief questionnaire about the event. However, informal feedback has been positive and several participating women, who were initially invited as a guest, went on to host their own session.

In summary, peer sessions with women can help to support information sharing about cardiovascular health, and the factors that are infrequently discussed during the clinical encounter, including stress, pregnancy, menopause, self-perception of health and experiences with the health system. These sessions, when co-facilitated by clinicians and designers trained in shared decision making can help to ensure that women receive evidence-based information in a way that allows reflection, discussion, and sharing of perspectives and experiences. The goal is that these sessions familiarize women with factors that are most relevant in their own cardiovascular health journey and help them prepare and feel more confident in their healthcare interactions.

## Data Availability

Data cannot be shared publicly because of potential for participant identification. Data are available in partnership with the investigators for researchers who meet the criteria for access to confidential data

## Acknowledgements

Funding for this project came from the Alpha Phi Foundation, Heart to Heart Grant.

## References

1. Agarwala A, Michos ED, Samad Z, Ballantyne CM, Virani SS. The Use of Sex-Specific Factors in the Assessment of Women’s Cardiovascular Risk. Circulation. 2020;141(7):592-599. doi:doi:10.1161/CIRCULATIONAHA.119.043429

2. Leifheit-Limson EC, D’Onofrio G, Daneshvar M, et al. Sex Differences in Cardiac Risk Factors, Perceived Risk, and Health Care Provider Discussion of Risk and Risk Modification Among Young Patients With Acute Myocardial Infarction: The VIRGO Study. Journal of the American College of Cardiology. 2015/11/03/ 2015;66(18):1949–1957. doi:10.1016/j.jacc.2015.08.859

3. Breslin M, Mullan RJ, Montori VM. The design of a decision aid about diabetes medications for use during the consultation with patients with type 2 diabetes. Patient Educ Couns. Dec 2008;73(3):465–72. doi:10.1016/j.pec.2008.07.024

4. Matlock DD, Spatz ES. Design and testing of tools for shared decision making. Circ Cardiovasc Qual Outcomes. May 2014;7(3):487–92. doi:10.1161/circoutcomes.113.000289

5. Montori VM, Breslin M, Maleska M, Weymiller AJ. Creating a conversation: insights from the development of a decision aid. PLoS Med. Aug 2007;4(8):e233. doi:10.1371/journal.pmed.0040233

6. Mullan RJ, Montori VM, Shah ND, et al. The diabetes mellitus medication choice decision aid: a randomized trial. Arch Intern Med. Sep 28 2009;169(17):1560–8. doi:10.1001/archinternmed.2009.293

7. Jhaveri A, Sibley RA, Spatz ES, Dodson J. Aspirin, Statins, and Primary Prevention: Opportunities for Shared Decision Making in the Face of Uncertainty. Curr Cardiol Rep. May 7 2021;23(6):67. doi:10.1007/s11886-021-01499-y

8. Weymiller AJ, Montori VM, Jones LA, et al. Helping patients with type 2 diabetes mellitus make treatment decisions: statin choice randomized trial. Arch Intern Med. May 28 2007;167(10):1076–82. doi:10.1001/archinte.167.10.1076

9. Mann DM, Ponieman D, Montori VM, Arciniega J, McGinn T. The Statin Choice decision aid in primary care: a randomized trial. Patient Educ Couns. Jul 2010;80(1):138–40. doi:10.1016/j.pec.2009.10.008

10. Perestelo-Pérez L, Rivero-Santana A, Boronat M, et al. Effect of the statin choice encounter decision aid in Spanish patients with type 2 diabetes: A randomized trial. Patient Educ Couns. Feb 2016;99(2):295–9. doi:10.1016/j.pec.2015.08.032

11. Kasl SV, Cobb S. Health behavior, illness behavior, and sick role behavior. I. Health and illness behavior. Arch Environ Health. Feb 1966;12(2):246–66. doi:10.1080/00039896.1966.10664365

